# Decomposing Heterogeneity in Disease Progression Speeds and Pathways

**DOI:** 10.64898/2026.01.30.26345194

**Authors:** Yuichiro Yada, Honda Naoki, the Pooled Resource Open-Access ALS Clinical Trials Consortium

**Affiliations:** Laboratory for Data-driven Biology, Nagoya University Graduate School of Medicine, 65, Tsurumai-cho, Showa-ku, Nagoya, Aichi, 466-8550, Japan; Laboratory of Data-driven Biology, Graduate School of Integrated Sciences for Life, Hiroshima University, Kagamiyama, Higashi-hiroshima, Hiroshima, 739-8526, Japan; Center for One Medicine Innovative Translational Research (COMIT), Nagoya University, 65, Tsurumai-cho, Showa-ku, Nagoya, Aichi, 466-8550, Japan

**Author notes:** Corresponding author: Yuichiro Yada, Address: Graduate School of Medicine, 65, Tsurumai-cho, Showa-ku, Nagoya, Aichi, 466-8550, Japan. Co-Corresponding author: Honda Naoki, Address: Graduate School of Medicine, 65, Tsurumai-cho, Showa-ku, Nagoya, Aichi, 466-8550, Japan. Data used in the preparation of this article were obtained from the Pooled Resource Open-Access ALS Clinical Trials (PRO-ACT) Database. As such, the following organizations and individuals within the PRO-ACT Consortium contributed to the design and implementation of the PRO-ACT Database and/or provided data, but did not participate in the analysis of the data or the writing of this report: - ALS Therapy Alliance - Cytokinetics, Inc. - Amylyx Pharmaceuticals, Inc. - Knopp Biosciences - Neuraltus Pharmaceuticals, Inc. - Neurological Clinical Research Institute, MGH - Northeast ALS Consortium - Novartis - Orion Corporation - Prize4Life Israel - Regeneron Pharmaceuticals, Inc. - Sanofi - Teva Pharmaceutical Industries, Ltd. - The ALS Association - The Sean M. Healey & AMG Center for ALS at Massachusetts General Hospital.

## Abstract

Understanding why patients with the same diagnosis exhibit markedly different disease progression—some progressing rapidly, others slowly, and through distinct symptom patterns—remains a major challenge in medicine. Here, we developed a machine learning framework called DiSPAH (Disease-progression Speed and Pathway Analysis based on a Hidden Markov model) to estimate both the pathway and speed of disease progression in individual patients. DiSPAH models disease progression as transitions of latent states evolving over continuous time with a patient-specific progression speed. We applied DiSPAH to longitudinal clinical scores from an amyotrophic lateral sclerosis (ALS) cohort and successfully inferred each patient’s hidden disease trajectory and progression speed. These individualized dynamics were significantly associated with baseline clinical features and enabled prediction of future disease course from data available at the first clinical visit. Our results highlight that jointly modeling progression pathway and speed improves prediction of heterogeneous disease courses, offering a powerful tool for personalized care and research in ALS and other chronic conditions.

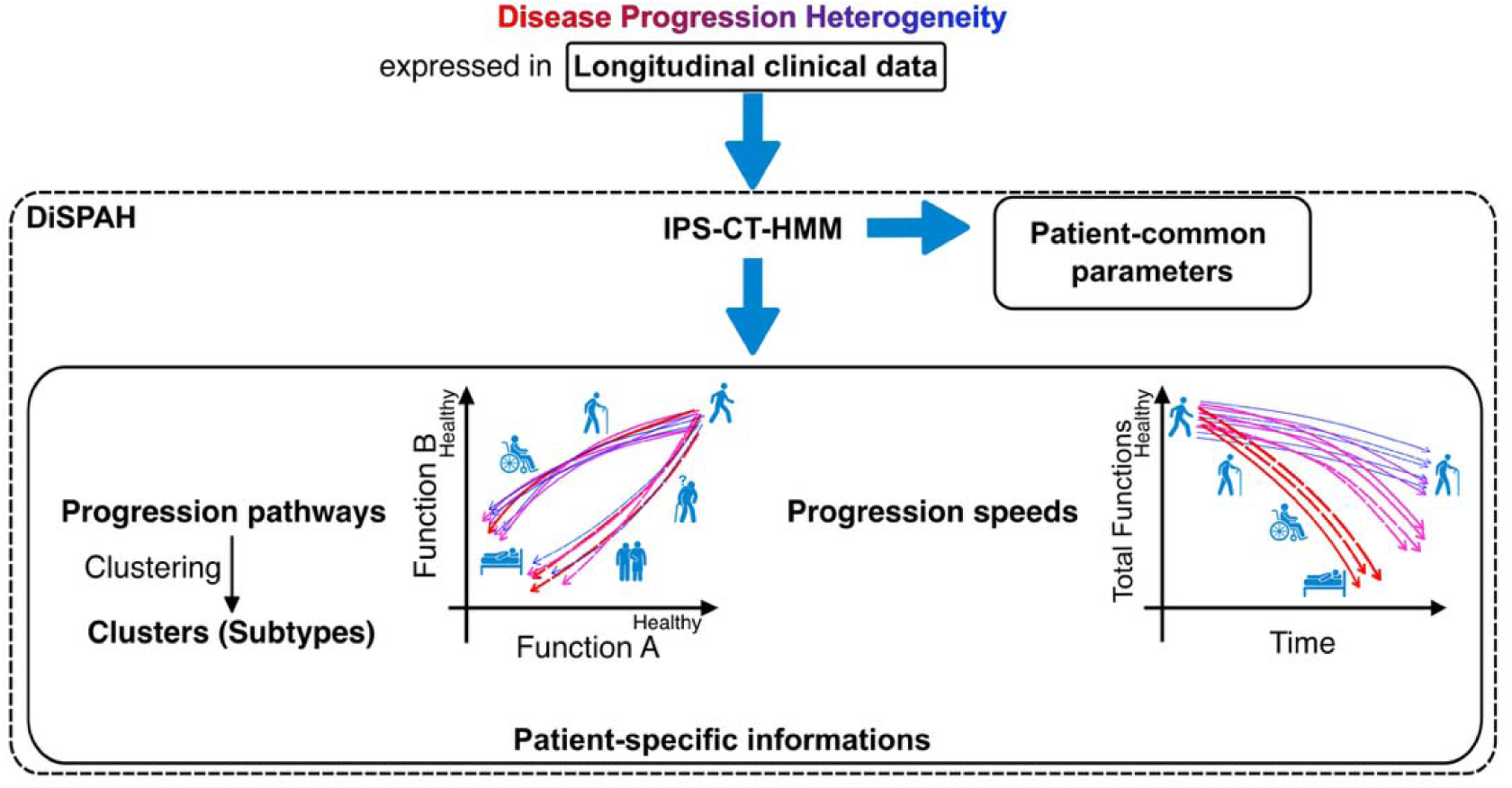
Graphical Abstract: Schematic illustration of the computational framework proposed in this paper

## Introduction

Why do patients with the same diagnosis follow drastically different courses—some deteriorating rapidly, others slowly, and often through entirely different patterns of symptom progression? Understanding and predicting the individual course of disease progression remains one of the most challenging problems in medicine^1–9^. In chronic and currently incurable diseases such as amyotrophic lateral sclerosis (ALS), this variability is not only clinically frustrating but also impedes effective treatment planning, clinical trial design, and patient counseling^8,10,11^.

To tackle this challenge, it is critical to decompose the heterogeneity of disease progression into two orthogonal dimensions (Fig. 1a). The first is the progression pathway, which describes which functions deteriorate first and in what order. The second is the progression speed, which quantifies how quickly a patient moves through the stages of disease. These two axes of heterogeneity may arise from different biological mechanisms, but most previous studies have not clearly distinguished them. A framework that can disentangle these aspects is essential for capturing the full complexity of disease dynamics at the individual level.

**Figure 1:**
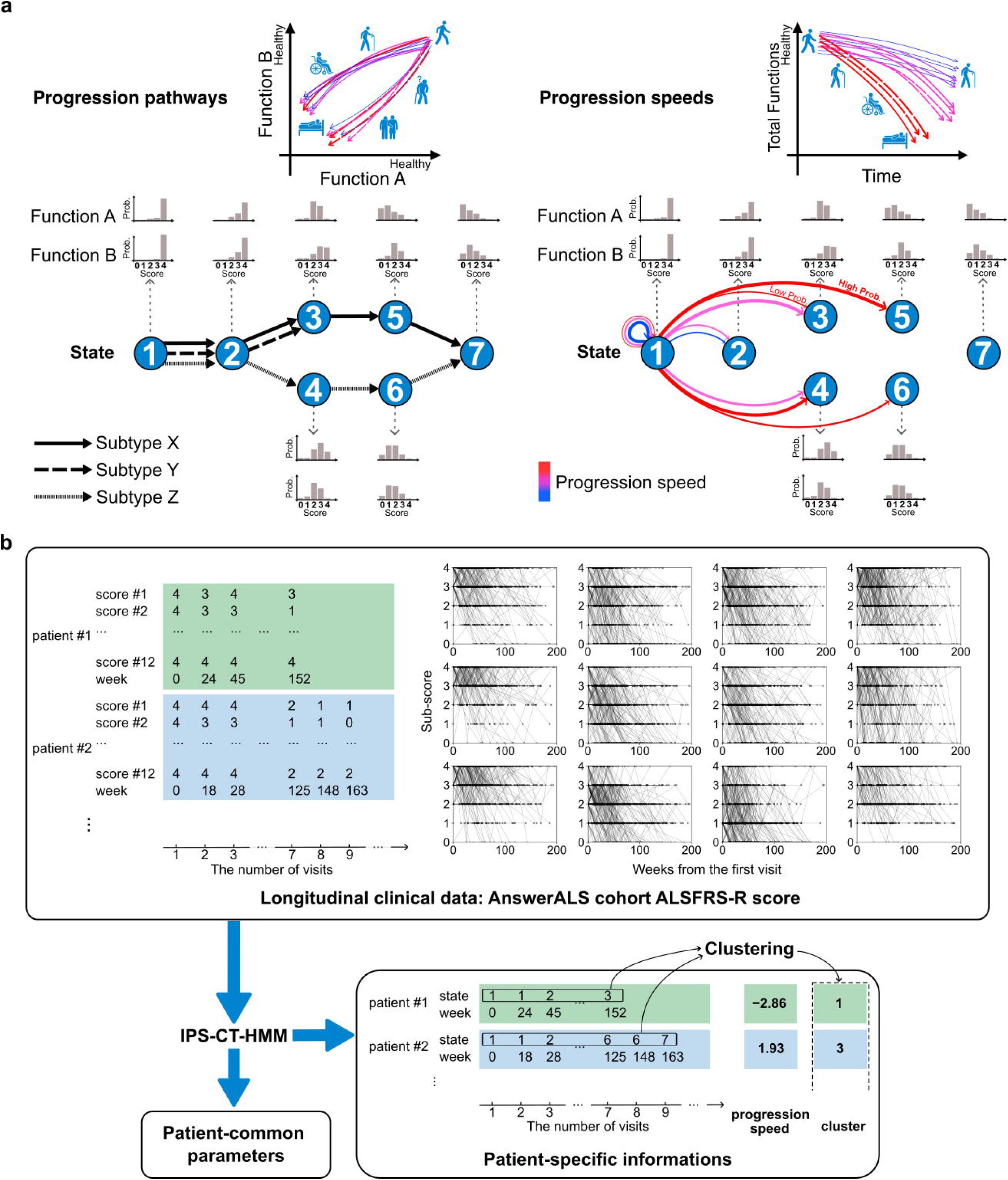
The concept of the present study. **a**, The concept of the present study and the proposed model. The progression of chronic diseases, including neurodegenerative diseases, varies greatly among individuals. The heterogeneity of disease progression includes not only heterogeneity in the progression pathway, i.e., which functions deteriorate preferentially, but also heterogeneity in the speed of progression. The proposed individual-progression-speed continuous-time hidden Markov model can deal both heterogeneities. **b**, Schematic diagram of the analysis flow with disease-progression speed and pathway analysis based on hidden Markov model (DiSPAH). Clinical longitudinal data, irregularly collected at the different number of time points for each patient, is input into IPS-CT-HMM for learning. The ALS Functional Rating Scale for the Residual Status (ALSFRS-R) is a clinical functional score for ALS consisting of 12 questions scored from 0 to 4 points, divided into four domains. Each column represents questions from a different domain. IPS-CT-HMM estimates patient-common parameters defining potential disease progression states, along with patient-specific patient-specific trajectories of disease progression states and disease progression speeds. By clustering the estimated trajectories, patient clusters (subtypes) with similar trajectories are obtained.

ALS exemplifies a disease in which heterogeneity in both progression pathway and speed significantly contributes to clinical variability^11,12^. It is a neurodegenerative disorder in which patients gradually lose control of voluntary muscles due to the degeneration of motor neurons in the central nervous system^13^. While progression is typically rapid with an average life expectancy of approximately 3 years after diagnosis^12^, some patients survive for more than 10 years^14,15^. Long-term survival is more frequently observed in younger-onset and limb-onset patients^14,16^. ALS also shows phenotypic heterogeneity, including variation in symptom onset site and the pattern of functional decline^17–19^. Currently, no radical cure exists, and only a few drugs are approved that slightly slow disease progression. This wide variation complicates prognosis and hinders therapeutic development. A framework that separately models progression pathway and speed is thus essential for advancing ALS research and personalized care.

In this study, we developed a machine learning framework called DiSPAH (Disease-progression Speed and Pathway Analysis based on a Hidden Markov model) to jointly estimate progression pathway and progression speed at the individual level. DiSPAH employs a continuous-time hidden Markov model (CT-HMM)^20–26^, in which clinical features—observed at irregular hospital visits—are probabilistically generated from latent disease states. Transitions between these latent states capture the diversity of progression pathways. To model individual differences in progression speed, we introduced a patient-specific speed parameter. We applied DiSPAH to longitudinal data from ALS cohort^27^ to estimate each patient’s hidden disease trajectory and progression speed. We further showed that these individualized dynamics were associated with baseline clinical features and that both progression speed and pathway could be predicted from data available at the first clinical visit. DiSPAH thus provides a powerful framework for uncovering heterogeneous disease progression across individuals.

## Results

### A framework to analyze heterogeneous disease progression

We have developed a machine learning framework for analyzing longitudinal clinical data from patients with diverse disease progression pathways and speeds, which we call DiSPAH (Disease-Progression Speed and Pathway Analysis based on a Hidden Markov model). DiSPAH is based on HMM to model how patients’ symptoms evolve over time, as recorded in longitudinal clinical data. Observed symptoms (e.g., clinical scores) are assumed to be probabilistically generated from underlying unobservable, latent disease states. These disease states are discrete and evolve over time through probabilistic transitions. To handle irregular timing of hospital visits, DiSPAH adopts a continuous-time HMM (CT-HMM), which describes the transition probability depending on time interval between observations. To reflect individual differences in disease progression speed, DiSPAH introduces a patient-specific speed parameter, which proportionally scales transition probability for each patient. This means that patients with larger progression speed parameters have a higher probability of transitioning to a worse state than those with smaller parameters (Figure 1a).

By learning the time series data of the clinical scores at various time points from a population of ALS patients, DiSPAH enables estimation of (i) a definition of the latent disease states that are represented by probabilistic distribution of clinical observations, (ii) individualized progression trajectories through the hidden disease states, and (iii) progression speeds for each patient. Furthermore, clustering the inferred trajectories allows the identification of patient subgroups with similar state transition patterns (i.e., disease progression pathways) (Fig. 1b). Detailed explanations of the proposed framework and the estimation algorithm are described in Methods.

### Application to longitudinal data from ALS patients

We applied the developed DiSPAH to the longitudinal ALSFRS-R scores from AnswerALS cohorts^27^. The ALSFRS-R score is a commonly used clinical scale for evaluating the functions of ALS patients^28^. It consists of 12 questions in total, with three questions for each of the four functional domains. Each question is scored on a scale of 0 to 4 (Fig. 1b). For this analysis, we included only patients with at least four visits to the hospital. Given the well-documented heterogeneity in progression pathways and speeds by site of disease onset^13–16^, we restricted our analysis to patients presenting with limb-onset ALS. A total of 264 patients met these criteria and are included in the analysis. The EM algorithm was used to learn the ALSFRS-R time-series data (Supplementary Fig. 1), estimating the following three model parameters common to all patients: the emission probability matrix, which describes the probability of observing a certain score on a given question in each hidden state (Fig. 2a, left), the transition rate matrix, which indicates transition probability from one hidden state to another (Fig. 2a, center), and the initial state probability, which is the probability of being in a certain hidden state at the start of tracking (Fig. 2a, right). Since the patient’s disease condition is expected to worsen monotonically over time in ALS or other neurodegenerative diseases, we assumed that transitions of disease state occur in a one-way fashion. From the ALSFRS-R time-series data, we estimated not only patient-common parameters, but also patient-specific disease progression speeds. The estimated progression speeds appeared correlated with the slope of the regression line to the total score of ALSFRS-R^27^, but a distinct difference between them was observed in a certain number of patients (Supplementary Fig. 2). Using the estimated parameters, we further inferred patient-specific trajectories of hidden states. By clustering these trajectories, we obtained six clusters of patients who showed similar disease progression pathways (Fig. 2b). While progression speeds tended to be localized within clusters, there was notable overlap in the distributions across clusters, suggesting that even patients with similar progression speeds may follow different latent-state progression pathways (Fig. 2c). This indicates that the proposed method can separately analyze the diversity of disease progression pathways and the diversity of progression speeds (Fig. 2d).

**Figure 2:**
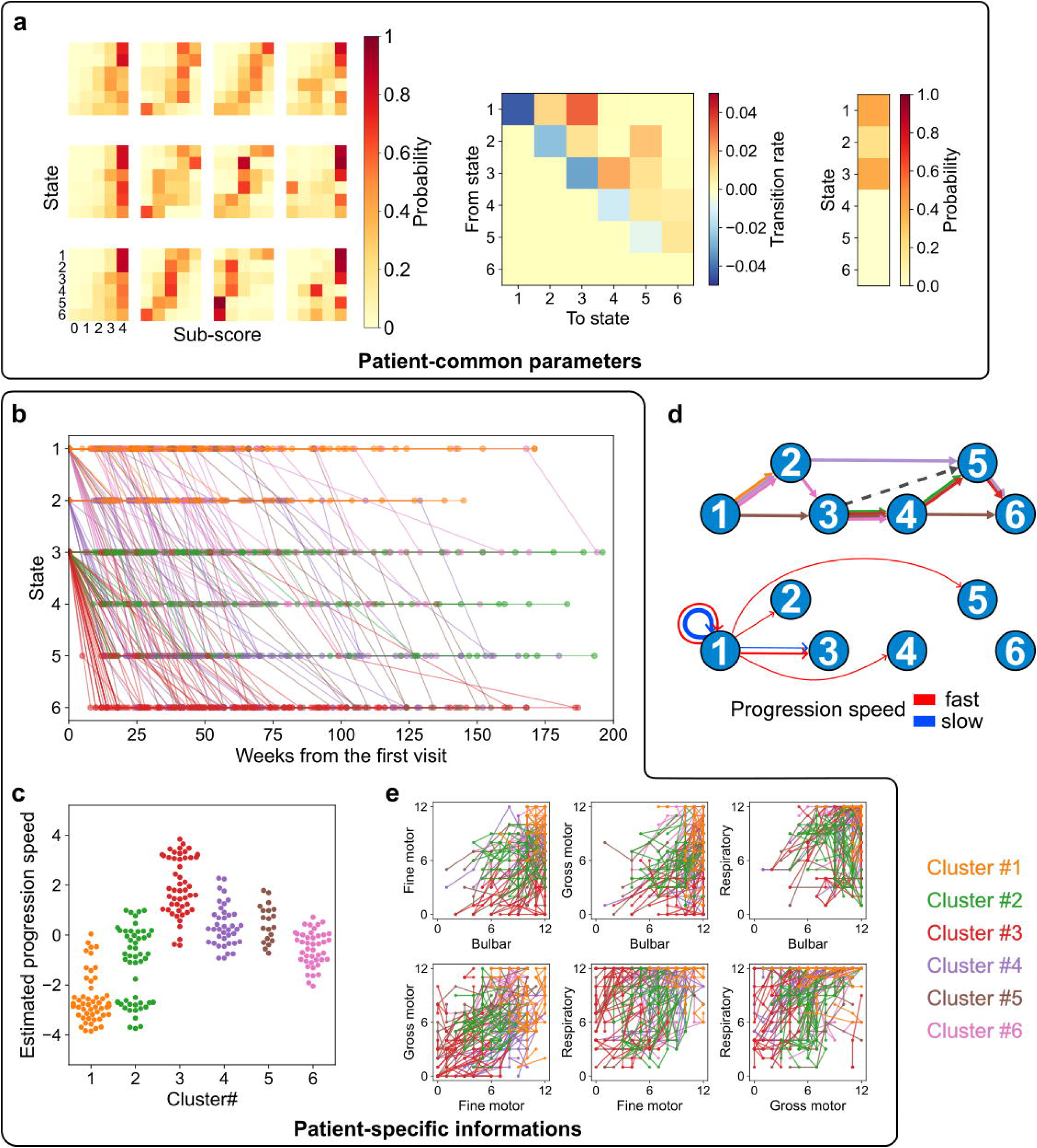
DiSPAH elucidated the heterogenous disease progression in AnswerALS cohort. **a**, Patients-common parameters estimated from the ALSFRS-R longitudinal data of AnswerALS cohorts. Emission probability matrices (left: the probability of observing a given score at each disease progression state), transition ratio matrix (center: the proportion of transitions from one state to another), and initial state probabilities (right: the probability of being in each disease progression state at the start) were estimated. The emission probability matrices include 12 matrices for each question. Each column represents questions from a different domain. **b**, The estimated patient-specific trajectories of disease progression states. **c**, The estimated patient-specific disease progression speeds. **d,** The schematic diagram of the estimated heterogeneous disease progression pathway (upper) and speed (bottom). In the pathway diagram, trajectories were drawn based on representative staying states within each cluster. A dashed line indicates a state transition that seems to be possible in the transition ratio matrix but is not included in the representative state transitions of the cluster. In the speed diagram, “fast” represents the probability of state transition over 5 weeks with, while “slow” a probability of 0.05 or higher are indicated by arrows. **e**, Patient-specific trajectories in the domain total scores of AnswerALS cohort patients. The domains are compared to show which domain deteriorates more rapidly. In **b**, **c**, **d**, **e**, each color represents a patient cluster obtained by clustering the estimated progression trajectories.

Each cluster exhibited characteristic disease progression pathways. Cluster #1 included patients with slow disease progression speed, remaining in stages 0 and 1. These patients showed gradual deterioration in motor function, but little decline in bulbar or respiratory function. Cluster #2 patients had motor function decline from the start of follow-up, but the progression speed was similar to that of Cluster #1, and they did not reach a state of complete functional deterioration. In contrast, Cluster #3 patients began at a stage similar to those in Cluster #2 but exhibited rapid progression, with deterioration predominantly in motor function, eventually reaching hidden state 5 (i.e., final ALS state). Cluster #4 was characterized by an unusual pattern in which gross motor function deteriorated earlier than fine motor function. Cluster #5 showed a progression speed comparable to Cluster #4 but with the opposite pattern: fine motor function declined earlier than gross motor function. Cluster #6 began with functional levels similar to Cluster #1 but experienced a slightly faster and more generalized functional decline overall (Fig. 2e, Supplementary Fig. 3).

### Verification of the estimated disease states and clusters using a larger cohort

To assess the universality of the probabilistic definition of ALS progression states and progression pathway clusters obtained from the AnswerALS cohort, we applied DiSPAH to the larger PRO-ACT cohort data (Fig. 3a). Following the same criteria used in AnswerALS to select patients, 2,565 individuals were included in the analysis. When training the IPS-CT-HMM model in the PRO-ACT cohort data, we incorporated patient-common parameters, i.e. the emission probability matrix, the transition rate matrix and the initial state probability, previously estimated from the AnswerALS cohort as prior information and used them without updating, allowing the model to infer only patient-specific information for the PRO-ACT data. Furthermore, for each patient in the PRO-ACT cohort, we compared the estimated state transitions with those of AnswerALS cohort patients and classified them into the AnswerALS cohort patient cluster with the closest sequence of staying states. As a result, we obtained that the distribution of functional decline pathways (Fig. 3b) and disease progression speeds (Fig. 3c) among patients within each cluster in the PRO-ACT cohort closely resembled those in the AnswerALS cohort. This consistency suggests that our proposed DiSPAH framework captures cohort-invariant features of ALS progression, rather than cohort-specific ones. However, one notable exception was observed: patients in Cluster #2 of the AnswerALS cohort showed a decline in respiratory function, whereas such pattern was not observed in the corresponding cluster of PRO-ACT cohort (Fig. 3b, Supplementary Fig. 4).

**Figure 3:**
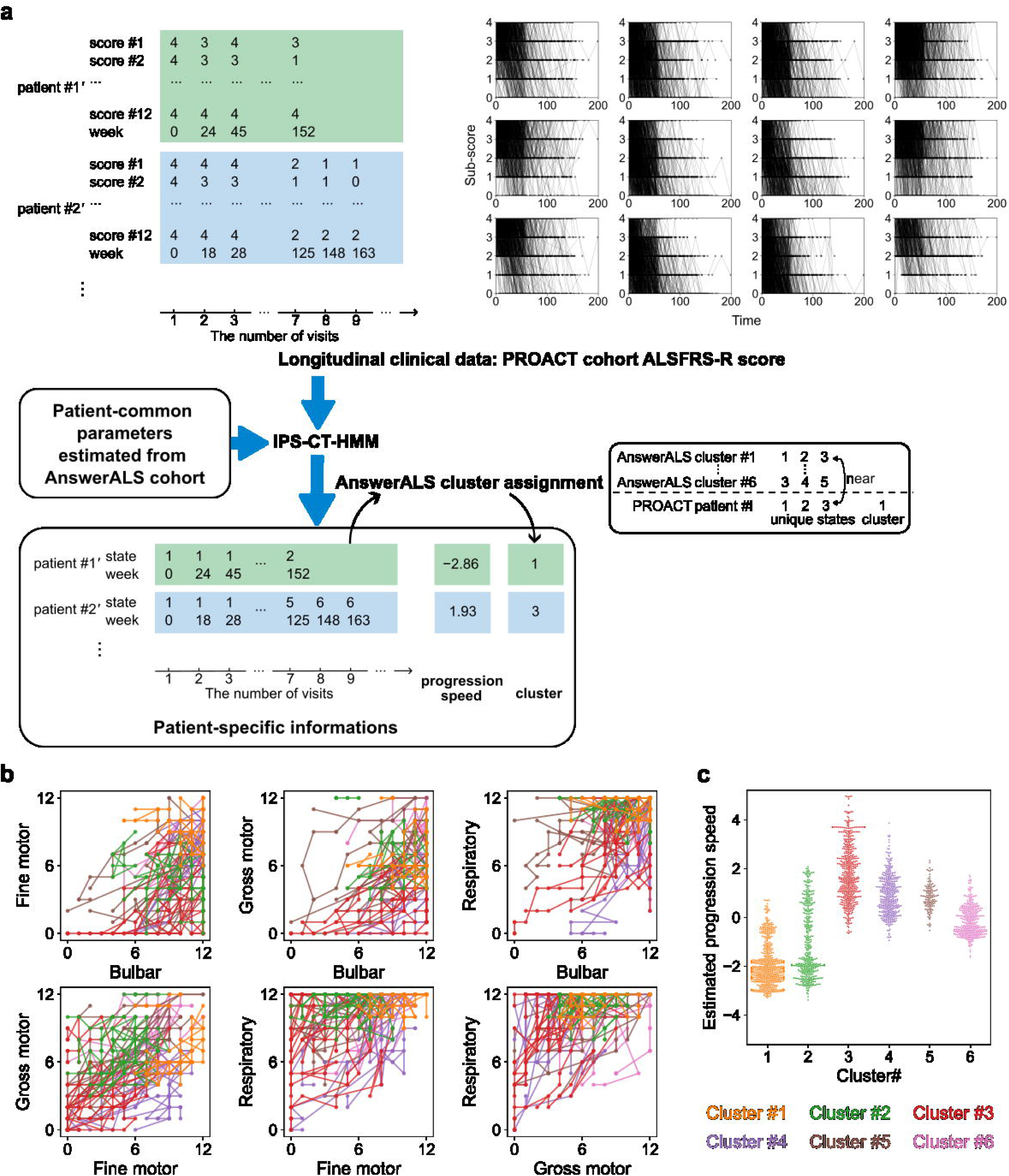
Verification of the elucidated heterogeneous disease progression in a larger cohort. **a**, Schematic diagram of the analysis flow of PRO-ACT cohort data using disease progression states estimated from AnswerALS cohort data. Longitudinal ALSFRS-R data is learned using IPS-CT-HMM. At this stage, patient-common parameters obtained from the AnswerALS data are provided to the model as prior information, and patient-specific progression trajectories and progression speeds are estimated. PRO-ACT cohort patients are classified into respective clusters by referencing the representative disease progression states of each cluster obtained from AnswerALS data. **b**, Patient-specific trajectories in the domain total scores of 20 PRO-ACT cohort patients randomly selected from each cluster. **c**, Distribution of the estimated patient-specific progression speeds for each cluster.

### Association between estimated progression speed and ALS-related genetic information

We next investigated the association between the estimated disease progression speed and known ALS-related genetic mutations. The AnswerALS cohort data contains information on these mutations identified in patients (Fig. 4a). Among these, C9orf72, ATXN2, and SOD1 mutations were identified in a sufficient number of patients and were therefore selected for further analysis. We performed a multivariate regression analysis to predict the estimated progression speed by the presence and absence of each gene mutations as explanatory variables, and also covariates of gender, age at onset, and prior exposure to riluzole^16,29^ (Supplementary Fig. 5a, b, c). The analysis revealed that only the C9orf72 mutation had a statistically significant positive regression coefficient (Fig. 4b), meaning that patients with the C9orf72 mutation exhibit faster disease progression compared to those without it. The result finding is consistent with the previous report linking C9orf72 mutation to shorter survival duration in ALS^30,31^. The patients who have SOD1 mutation appeared to have relatively slow progression, but no statistically significant differences were observed. No significant associations were also found in the patients with ATXN2 mutations (Supplementary Fig. 5d, e).

**Figure 4:**
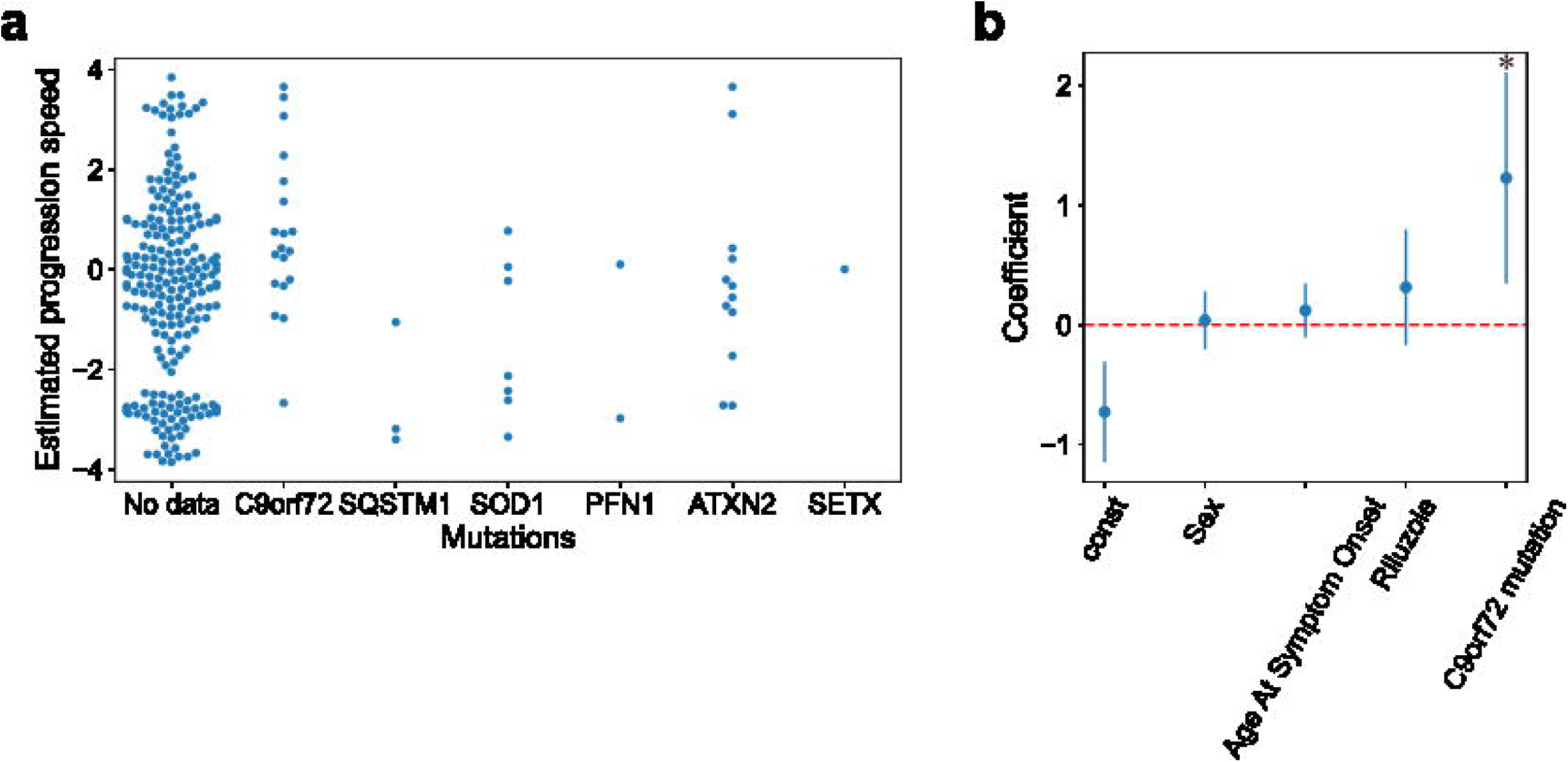
Association between estimated disease progression speed and ALS-related mutations. **a**, Relationship between ALS-related gene mutations and estimated progression speeds. **b**, Coefficients estimated by multivariate regression with the presence or absence of C9orf72 mutation as the main explanatory variable and the estimated progression speeds as the target variable, adjusted for sex, age at symptom onset, and riluzole use. The error bars indicate 95% confidence intervals.

### Discovering biological associations of disease progression speed using patient iPSC-derived motor neurons

To explore biological factors associated with the progression speed of ALS, we evaluated the relationship between genome-wide gene expression in motor neurons derived from patients. In the Answer ALS dataset, induced pluripotent stem cells (iPSCs) were established from patients, differentiated into motor neurons (iPSC-MNs), and subjected to transcriptome and proteome data^27^ (Fig. 5a).

**Figure 5:**
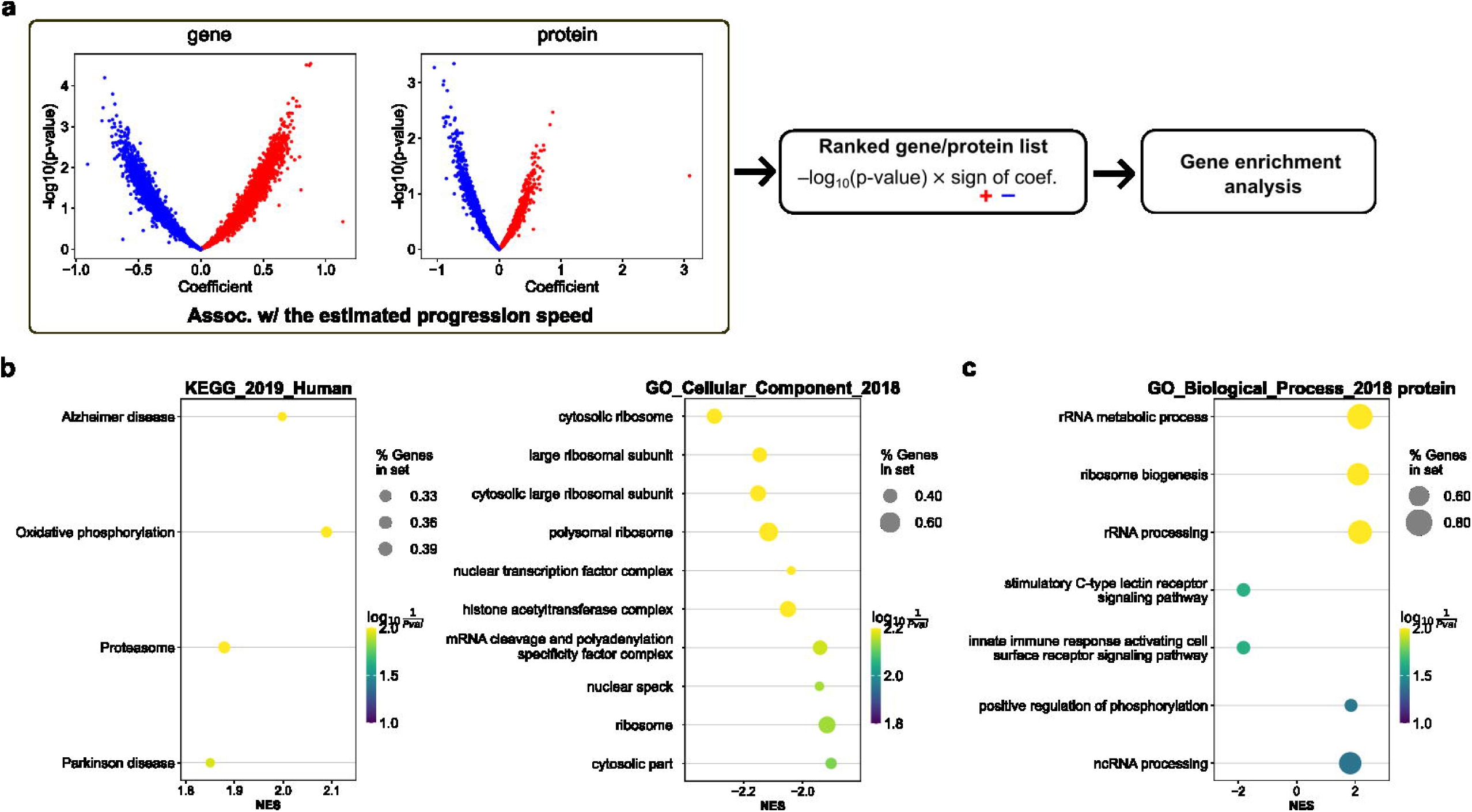
Analysis of the association between gene/protein expression in patient-derived iPSC-MNs and estimated disease progression speed. **a**, The workflow in Gene enrichment analysis. First, perform multivariate regression with specific gene/protein expression levels as the primary explanatory variables and estimated progression speeds as the target variables, then rank the genes/proteins based on the p-values and the estimated coefficient signs. Use this ranking to perform GSEA analysis. **b**, Examples of gene groups exhibiting significant fluctuations in relation to estimated progression speed, obtained from GSEA analysis in the transcriptome. **c**, Those obtained from the proteome.

To examine the association between the DiSPAH-estimated disease progression speed and individual gene expression in the motor neurons, we performed multivariate regression analysis for each gene, in which disease progression speed was set as the target dependent variable, and expression level of each gene as explanatory variable. Sex, age at onset, and riluzole exposure status were included as covariates in all models. No single gene was identified as significantly influencing disease progression speed (Fig. 5a). To explore broader functional trends, we ranked genes based on p-values and regression positive or negative regression coefficients and conducted gene enrichment analysis using GSEA^32,33^. In GSEA, the analysis targeted KEGG pathways and Gene Ontology (GO) categories: Biological Process (BP), Cellular Component (CC), and Molecular Function (MF). In the KEGG pathway analysis, significant upregulation was observed in pathways related to neurodegenerative diseases such as Alzheimer’s disease and Parkinson’s disease, as well as in pathways related to oxidative phosphorylation and proteasome function. In GO CC analysis, we observed downregulation of gene clusters associated with mature ribosomes, such as polysomes (Fig. 5b, Supplementary Fig.6a, b).

We also applied a same multivariate regression analysis to the proteomic data, evaluating the association between protein expression and the DiSPAH-estimated disease progression speed. As with the transcriptomic data, no significant association with a single protein was observed (Fig. 5a). However, GSEA analysis revealed upregulation of protein groups related to ribosome biogenesis in GO BP category (Fig. 5c, Supplementary Fig.6c).

The results suggest the following pathophysiological cascade: abnormal protein aggregation leads to the suppression of global translation, triggering a compensatory up-regulation of ribosome biogenesis, resulting in the accumulation of immature ribosomal precursors. In parallel, cells boost oxidative-phosphorylation pathways to secure the ATP required for proteasome-mediated clearance of mis-folded or surplus proteins. Mitochondria stressed under this high-flux state leak electrons, raising reactive-oxygen-species (ROS) levels. The combined burden of translation arrest, proteostasis overload, and oxidative stress is hypothesized to accelerate motor-neuron damage, thereby amplifying disease progression through a vicious cycle of cellular dysfunction.

### Prediction of disease progression speed and pathway based on data available at the initial time point

Finally, we evaluated the feasibility of predicting patient-specific progression speed and progression-trajectory cluster, which were estimated from longitudinal data by DiSPAH, based on information available at the start of follow-up. Specifically, we used the following features as input variables for prediction: (i) the sub-total score of ALSFRS-R for each functional domain (0-12 for Bulbar, Fine motor, Gross motor, and Respiratory functions), and (ii) the presence or absence of C9orf72 and SOD1 mutations, both of which were selected based on our multivariate genetic analyses and the GSEA results obtained from iPSC-MN omics data. These features were used as input variables, and the previously estimated progression speed or cluster labels were used as output variables. The relationships between inputs and outputs were learned by ridge regression (for progression speed regression) and support vector machine (SVM) with linear kernel (for cluster classification). To evaluate predictive performance, we applied leave-one-out cross-validation. In each iteration, the model was trained on a dataset excluding one patient, and the trained model was used to predict the progression speed or cluster label of the excluded patient. This process was repeated for all patients, and the overall prediction accuracy was evaluated (Fig. 6a).

**Figure 6:**
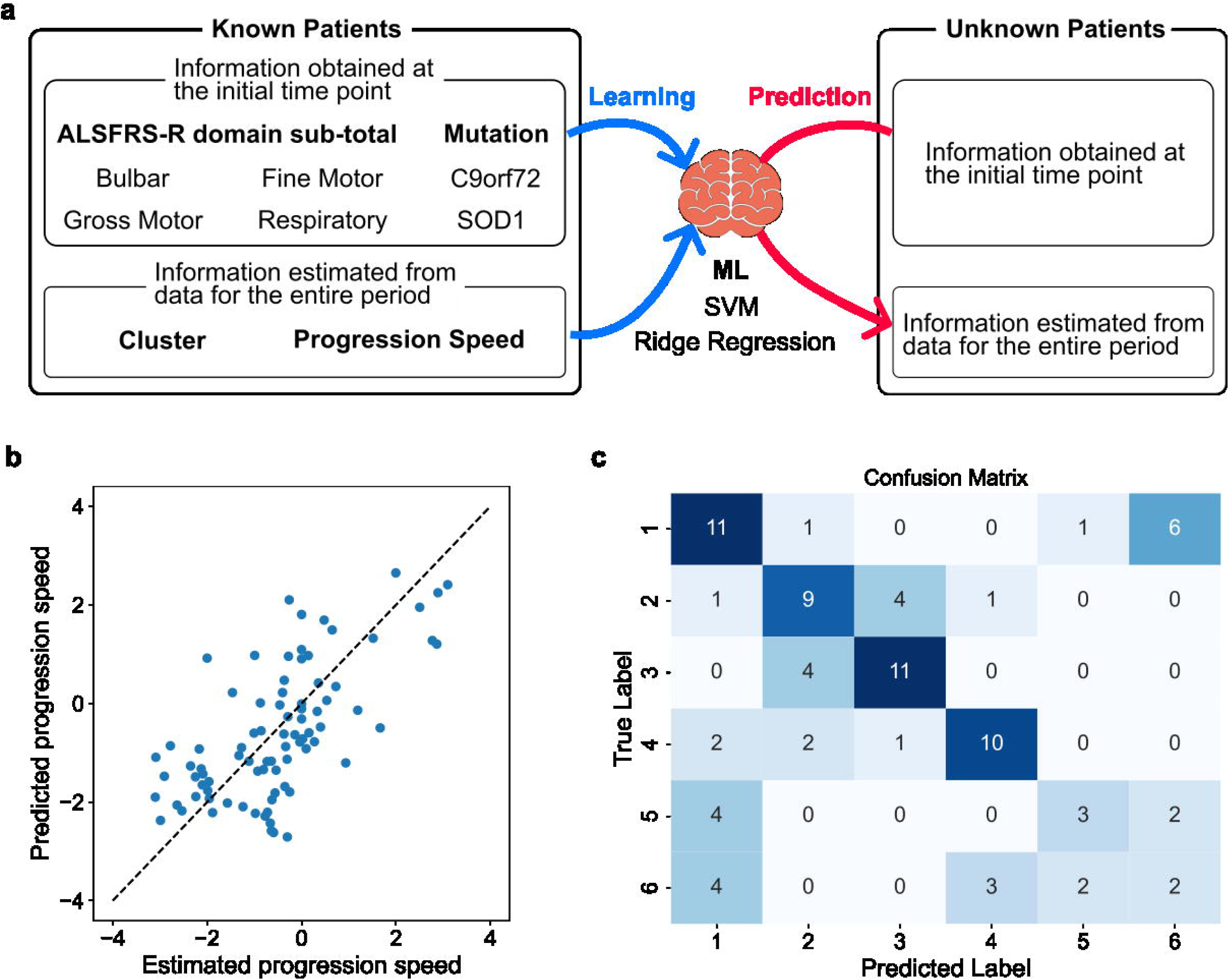
Prediction of disease progression speed and clusters based on information available at the start of tracking. **a**, Analysis flow for predicting progression clusters and progression speeds based on information available at the initial time point for the follow-up. **b**, The result for progression speed predictions. The progression speeds predicted from initial information and the progression speeds estimated from the whole period of data match on the dotted line. **c**, The result for progression cluster predictions.

The trained regression model showed reasonable performance in predicting progression speed, particularly for patients showing very slow and very fast estimated progression speeds. The mean squared error was 1.28, and the coefficient of determination was 0.34 (Fig. 6b). For progression cluster prediction, we also used the predicted progression speed as input for SVM in addition to the information available at the start of follow-up. As a result, we were able to correctly predict disease progression clusters with approximately 54.8% accuracy (Fig. 6c). Among the clusters, clusters #2 and #3, as well as clusters #5 and #6, showed relatively similar progression patterns. When these pairs of clusters were combined as broader clusters, the prediction accuracy improved to approximately 69.0%, indicating that the proposed method can effectively distinguish major progression trends (Fig. 6c). Gene expression levels in patient iPSC-derived motor neurons did not contribute to predicting progression speeds or progression pathway clusters (Supplementary Fig. 7). The results suggest that the proposed method may provide useful prognostic information at an early stage after onset.

## Discussion

We proposed DiSPAH, a modeling framework combining IPS-CT-HMM and trajectory clustering to dissect heterogeneity in ALS progression. Applied to the AnswerALS cohort, DiSPAH estimated latent disease states, individual progression speeds, and stratified patients into six distinct trajectory clusters. These findings generalized to the PRO-ACT cohort, supporting the model’s robustness. A C9orf72 mutation was linked to faster progression, and multi-omics analysis of iPSC-MNs revealed molecular signatures of disrupted proteostasis and oxidative stress in fast progressors. Moreover, progression speed and cluster identity could be partially predicted from initial clinical and genetic information, highlighting the potential of DiSPAH for early prognosis and personalized trial stratification in ALS.

Longitudinal tracking of clinical scores, biomarkers, and neuroimaging has enabled data-driven stratification of disease progression in ALS and other neurodegenerative disorders^8,34,35^. Recent machine learning-based approaches have identified subgroups of patients with distinct progression trajectories, shedding light on disease heterogeneity. In ALS, clustering of longitudinal ALSFRS-R data reveal heterogeneous progression subtypes^11^. A recent approaches applying Gaussian mixture processes^8^ or triclustering^11^ to longitudinal data from the ALSFRS-R revealed heterogeneous subtypes of progression. Similar machine learning approaches have been applied to other neurodegenerative diseases. In Alzheimer’s disease, subtypes have been identified using event-based models^4,36,37^ and Bayesian clustering on biomarker and brain imaging data^38^. In Parkinson’s disease, subtypes have been identified by latent time joint mixed-effects models and deep generative model-based clustering^39^ to clinical scores and biomarkers. Among these previous studies, subtypes exhibiting characteristic progression speeds have been reported^39^. Unlike these existing methods, the proposed method explicitly separates the estimation of progression speeds from the clustering of progression pathways, enabling the identification of factors influencing progression speeds. This offers a different perspective to previous methods, and may promote a more detailed understanding of disease mechanisms.

In this study, we defined disease progression speed as a parameter that modulates the probability of state transitions occurring over a given time interval in a continuous-time hidden Markov model (CT-HMM)^20,22^, and estimated it from clinical data with irregular observation intervals. A widely used method for assessing ALS progression is to just fit a straight line to the total ALSFRS-R score over time and use the slope as an index of disease speed^40,41^. This conventional linear-fit approach assumes that overall functional decline in ALS proceeds uniformly and linearly across all domains, which rarely reflect actual disease dynamics^8^. In practice, however, symptom often worsen sequentially in specific domains or involves multiple functional systems progressing at different times, resulting in non-linear decreases in the total score. Moreover, in actual data, clinical scores may occasionally improve, leading to incorrect estimates when using linear fitting. By contrast, our probabilistic model-based approach is better suited to capturing these real-world complexities. When comparing the disease progression speeds estimated by DiSPAH with the ALSFRS-R progression slope recorded in the database, we observed a moderate correlation. However, many patients showed different relative rankings between the two measures (Supplementary Fig. 2). In addition, The ALSFRS-R slope showed a biased distribution with some patients exhibiting positive rather than negative slopes, raising concerns about whether conventional ALSFRS-R slope reliably reflects the true progression speed. These results suggest that the progression speed estimated by our model has the potential to serve as more robust and interpretable alternative indicator to the conventional slope-based index.

We identified six states and six clusters representing distinct disease progression pathways in the analysis of limb-type ALS patients. These states and clusters captured key aspects of progression, including: (i) how rapidly bulbar and respiratory functions declined relative to motor deterioration, (ii) whether fine-motor or gross-motor abilities deteriorated first, and (iii) whether a given function was already impaired at baseline or became completely lost by the end of follow-up. Regarding the third point, if our modeling and clustering framework could incorporate information beyond the observation window, it may be possible to stratify patients more appropriately. Clinically, limb-onset ALS is often categorized into upper limb-predominant and lower limb-predominant types^10,42^. The order of fine and gross motor decline observed in our results may reflect these known clinical subtypes.

As a limitation of the proposed model, there is a complementary relationship between the speed parameter and the transition ratio matrix. Hence, the absolute value of the estimated progression speed may depend on the oveall scale of the transition rates. However, when applied to real-world clinical data, an absolute notion of progression speed does not exist and is meaningful only in relative terms. Accordingly, this study focused on estimating relative progression speed across patients. We estimated the progression speed using a prior Gaussian distribution with a mean of zero. The choice of the hyperparameters value require further investigation, and alternative methods such as the method of Lagrange multiplier can be considered to regularize estimation. A detailed examination of the constraint setting method and calculation method is a future task.

In the proposed method, clusters were identified by hierarchical clustering based on estimated patient-specific state trajectories. An alternative approach to stratifying disease progression in patients involves incorporating subtypes directly as hierarchical latent variables within the probabilistic model and estimating them jointly with other parameters^8^. While such methods are promising, as they allow for comprehensive model-based clustering, they rely on the assumption of the existence of discrete subtypes, which may obscure continuous spectrum-like heterogeneity. For this reason, we selected to use post-hoc clustering in this study. An alternative approach involving the hierarchical assumption of continuous “disease progression pathway” variables rather than discrete ones is also worth considering for future research.

Our multivariate analysis identifies the C9orf72 hexanucleotide repeat expansion as the only mutation in the Answer ALS cohort that independently accelerates the data-driven estimate of disease progression speed, consistent with prior reports of shortened survival in C9orf72-associated ALS^30,31^. Mechanistically, our GSEA of patient-derived iPSC-MNs revealed a coherent molecular signature implicating global translational inhibition at the center of fast progression. Meanwhile, SOD1 inclusions formed in patients with SOD1 mutations are independent of TDP-43^43^ and may not directly involve the translational machinery. The prognosis of SOD1 mutation patients varies depending on the mutation type, with D90A and H46R showing relatively mild clinical courses^44,45^, while the structurally unstable A4V mutation causes a severe ALS progression^46^. Due to the lack of information on mutation types, further investigation is necessary; however, the results from this study suggested that the progression speeds in SOD1 mutation patients were relatively slow, and direct ribosomal interference, which is present in C9orf72 and TDP-43 proteinopathies but rarely observed in most SOD1 variants, may accelerate disease progression. Additionally, proteomics analysis of iPSC-MNs revealed a tendency for proteins with negative effects on disease progression speed to outnumber those with positive effects. That is, lower expression levels of many proteins were associated with faster disease progression (Fig. 5a). The converging results of this study support a unified concept: the capacity of motor neurons to maintain protein synthesis balance under aggregation stress is a major determinant of ALS progression speed.

## Methods

### Cohorts and data pre-processing

The Answer ALS cohort^27^ was used to estimate disease progression states and clusters, while the larger PRO-ACT cohort was used to validate the estimated states and clusters. The Answer ALS cohort includes information on the presence or absence of ALS gene mutations in patients and omics data from iPSC-MNs derived from patients, which were utilized in the analysis. In this study, we focused exclusively on patients diagnosed with ALS and classified as Limb-onset type. In both cohorts, patients were included if they had four or more hospital visits, had ALSFRS-R scores available, and had a total ALSFRS-R score of 21 or higher at the start of follow-up. The PRO-ACT cohort includes patients with very long follow-up periods and a large number of visits, so we set the maximum number of follow-up visits to 20 and the maximum follow-up period to 200 weeks, and only patients meeting these criteria were included in the analysis. Basic characteristic values for each cohort are shown in Supplementary Table 1.

For the ALSFTS-R score, the value recorded for questions 5a and 5b was used as the score for question 5. In addition, patients evaluated by ALSFRS-R, not by ALSFRS were included in the analysis. When any one of the scores for each question was missing, it was assumed that there was no observation point. That is, each observation point was ensured to have exactly 12 scores.

The Answer ALS cohort data contain information about major ALS-associated gene variants. The presence of the mutations in patients was detected based on whole-genome sequencing (WGS) results, or through the record in “ClinReport Mutations Details,” or, for mutations involving expansion of repetitive sequences, through records of the number of repeat sequences. The repeat expansion mutations were defined as being present when they met the following criterion^47,48^:

The number of repeats in C9orf72 ≥100

The number of repeats in ATXN2 ≥ 27

### IPS-CT-HMM: individual-progression-speed continuous-time hidden Markov model

The ALS pathology’s progression and clinical observation are modeled with an individual-progression-speed continuous-time hidden Markov model (IPS-CT-HMM). We assume that a patient *i*(*i* = 1, …, *I*) has *n_i_*-th observation at times *t_ni_*. The time interval of the observation of patient *i* is hence,

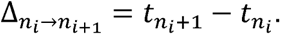

The clinical data observed from a patient *i*, at *n_i_* -th observation is denoted as 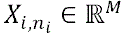, where *M*, is the dimension of observed data(*m*=1,2,…12). The hidden. Disease progression state of ALS progression at the *n_i_* -th observation is defined as 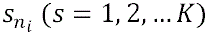. The probability of transition from state *k*, at time, *t_ni_* to state *j* at time *t_ni_*+1 is defined with the matrix exponential as

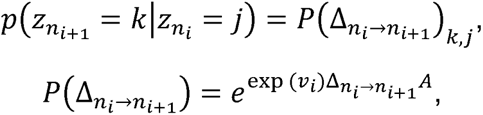

where *A* is a state transition rate matrix and *v_i_* denotes a patient-specific progression-speed parameter, whose exponential scales the transition frequency for that patient. We assumed that *v_i_* follows a normal distribution with a mean of zero

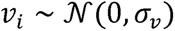

The diagonal elements of *A*, are defined as 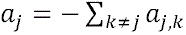, which means that the summation of elements of *A* in each row equals zero. In neurodegenerative diseases, it is currently impossible for disease progression to be reversed, i.e., for the loss of nerve cells to be improved. Therefore, we assumed that the progression state would not return to its original state. In other words, the state transition ratio matrix was set as an upper triangular matrix. In addition, since only patients who did not show a significant decline in ALSFRS-R at the start of follow-up were included, the initial state at that point was assumed to be either state 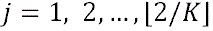.

Derivation of the EM algorithm of CT-HMM was described in the past literature^20,49^. In an IPS-CT-HMM case, the complete log-likelihood is formulated as follows:

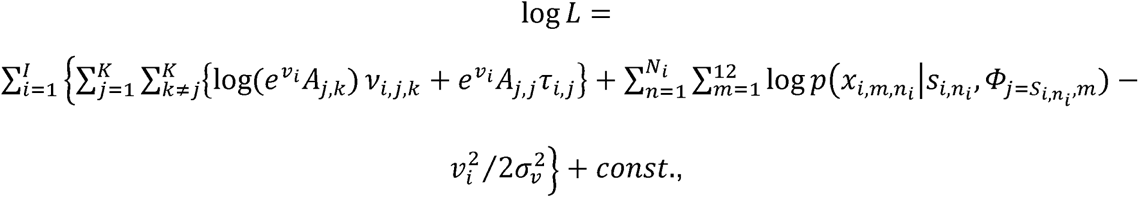

where *v_i,j,k_* is the times of individual *i*‘s transition from state *j* to state *k*, and *τ_i,j_* is the individual *i*‘s staying times at state 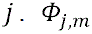 is an 5-dimensional (*l*=0,1,…4) categorical distribution representing the probability distribution of the ALSFRS-R *m*-th score at state *j*. Thus, the expected complete log-likelihood is as follows:

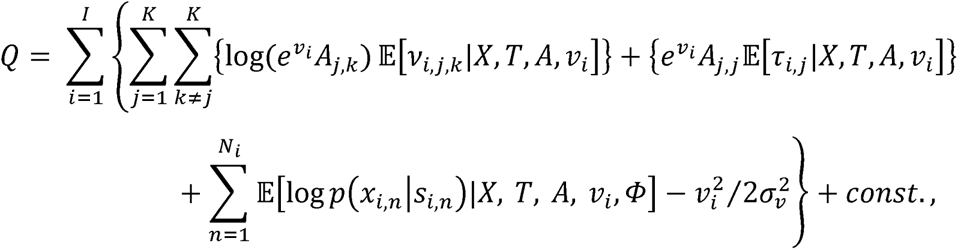

where the calculation methods for the expectation of *v_i,j,k_* and *τ_i,j_* were based on the methods described in the original paper on the CT-HMM algorithm^20^ (E-step).

In the M-step, the parameters were updated according to the following equations, which are the partial derivative of the expected complete log-likelihood:

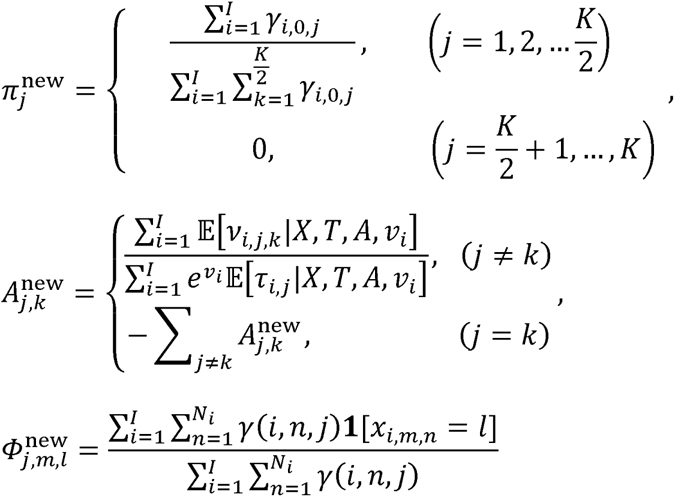

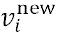 can be obtained by numerically solving the following equation. In this study, we used the BFGS algorithm to calculate the solution.

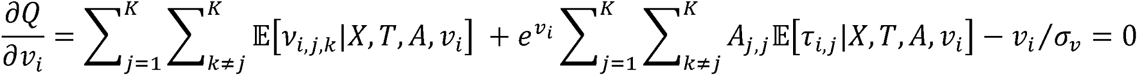

The patient-specific sequence of disease progression states for each patient 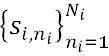 were estimated by using Viterbi algorithm. The number of the hidden disease states were determined to maximize likelihood via cross-validation^5^.

### Clustering of the trajectories of the disease progression states

By clustering the estimated trajectories of disease progression states, we obtained clusters of patients showing similar disease progression pathways. First, the estimated state series included information on the time when observations corresponding to the estimated states were made, but since we focused only on the pathways, we excluded time-related information. We also removed duplicate states from the state series and obtained a unique set of states for each patient. Next, we defined the distance between states using emission probability distributions. Specifically, we defined the Wasserstein distance between the emission probability distribution of each score in a given state and the emission probability distribution in another state as the distance between those states.

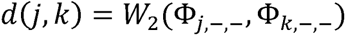

Using this definition of distance between states, we calculated the dissimilarity between sequences as Dynamic Time Warping (DTW) cost and defined the similarity of sequences as the negative value of DTW cost. Then, we performed hierarchical clustering using the Ward’s method to obtain patient clusters. The number of clusters was set to be sufficient for identifying clusters with different state transition branching in the estimated transition ratio matrix.

### Estimation for the PRO-ACT cohort patients using the results from the Answer ALS cohort

The parameters common for all patients, which include the state transition ratio matrix, the probability of each sub-score occurring in each state, and the initial state probability, were provided to the IPS-CT-HMM as known values, estimated from the AnswerALS patient data. Then, PRO-ACT cohort patient data was provided to estimate the disease progression speed and the trajectory of disease progression states specific to each PRO-ACT cohort patient.

Cluster assignment for PRO-ACT cohort patients was performed based on disease state trajectories, where clusters derived from AnswerALS cohort patient data were assigned. The disease state trajectories estimated for PRO-ACT cohort patients were compared to those of AnswerALS cohort patients. For each combination, the DTW cost was calculated as was done during the AnswerALS clustering. Subsequently, the AnswerALS patient cluster exhibiting the closest disease state transition was assigned as the disease progression cluster for each PRO-ACT cohort patient.

### Association analysis of genetic mutations with estimated disease progression speed

We modeled estimated progression speed as the dependent variable using ordinary least squares (OLS) regression implemented in Python. The primary explanatory variable was the presence of the genetic mutation of interest (no = 0, yes = 1). Covariates were sex (female = 0, male = 1), age at symptom onset (normalized to have a mean of 0 and a variance of 1) and history of riluzole use (no = 0, yes = 1). The model was

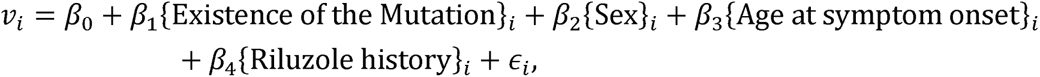

With *∊_i_* assumed i.i.d. with mean zero. Patients with any missing values in model variables were excluded. Our primary parameter of interest was *β*_1_, the coefficient on the mutation indicator. Two-sided t-tests at α=0.05 were used for inference; effects were deemed statistically significant when the two-sided 95% confidence interval for *β*_1_ excluded zero. A positive *β*_1_ indicates higher estimated progression speed in carriers relative to non-carriers. Model assumptions were evaluated by visual inspection of residual-versus-fitted plots and normal Q–Q plots to assess linearity, homoscedasticity, and approximate normality of residuals.

### Discovery of biological association of the disease progression speed

We obtained transcriptome and proteome data obtained from motor neurons differentiated from patient iPS cells from the AnswerALS database. First, we perform multivariate regression, where estimated progression speed as the dependent variable and the expression of a gene/protein as the main explanatory variable with covariates including sex, age of symptom onset and the existence of C9orf72 mutation. Next, we ordered genes/proteins based on their – log_10_ (p-value) from the multivariate regression results and performed gene enrichment analysis (GSEA). Here, the order was arranged by dividing the genes/proteins into those with positive effects on expression levels and those with negative effects on estimated progression speed, based on the multivariate regression results. Genes/proteins with positive effects were sorted in descending order by -log_10_ (p-value). Subsequently, genes/proteins with negative effects were sorted in ascending order by - log_10_ (p-value). GSEA was performed using pyGSEA. The gene sets used in the GSEA were “KEGG_2019_Human”, “MSigDB_Hallmark_2020”, “GO_Biological_Process_2018”, ‘GO_Molecular_Function_2018’, and “GO_Cellular_Component_2018”. The minimum and maximum size of the elements in the gene set was set as 15 and 1000, respectively, and the number of permutation times was set as 1000 times.

### Prediction of the disease progression speeds and progression clusters

We evaluated whether the disease progression speed and progression cluster of patients could be predicted from clinical information at the start of tracking. The AnswerALS dataset was separated into a training dataset that was used to train machine learning models and test data that was used to evaluate the performance of the trained machine learning models. The test dataset contained data of patients who do not belong to the training dataset. Cross-validation was performed in the leave-one-out manner. The assignment of clusters to patients within each cross-validation set was performed by defining characteristic residence states for clusters derived from the full AnswerALS cohort data. We estimated the state trajectories of patients within each cross-validation set and assigned the cluster whose characteristic staying state most closely matched their estimated disease state trajectory.

For the predictors, we used sex, age at symptom onset, the presence of the genetic mutations (C9orf72 and SOD1) and the sum of the scores for each of the four domains of the ALSFRS-R. When the analysis was configured to include omics features, the principal components of gene/protein expression levels were added to the pool of predictors.

To predict the estimated progression speed from the constructed predictors, we fitted an L2-regularized linear model (a Ridge regressor). the regularization strength to minimize mean squared error was selected by 5-fold cross-validation within the training set using a grid. The best model was refit on the full training set and used to generate predictions for the held-out test set.

We also predicted the categorical cluster label using a linear-kernel support vector machine. To capture the joint information in clinical features and disease speed, the classifier’s input comprised the same predictor variables used for predicting progression speeds plus a single additional feature: the progression speed estimated from whole observations for training and the predicted progression speed from the Ridge regressor for test. The regularization parameter was tuned by 5-fold cross-validation over with accuracy as the scoring metric, where class imbalance was adjusted. The selected SVM was then applied to the held-out test set to obtain predicted cluster labels.

## Data availability

Clinical data and iPSC-MN omics data from AnswerALS cohort are available for download from the Answer ALS data portal (https://dataportal.answerals.org). Clinical data from PRO-ACT cohort are available for download from the PRO-ACT database (https://ncri1.partners.org/ProACT/Home/Index).

## Code availability

All code used in the current study is available on the GitHub repository (https://github.com/yadapo/DiSPAH).

## Acknowledgements

Part of the data used in the preparation of this article were obtained from the ANSWER ALS Data Portal (AALS-01184). For up-to-date information on the study, visit https://dataportal.answerals.org.

Part of the clinical data used in the preparation of this article were obtained from the Answer ALS Foundation Program, ‘Answer ALS’. For up-to-date information on the program, visit https://www.answerals.org.

Part of the data used in the preparation of this article were obtained from the Pooled Resource Open-Access ALS Clinical Trials (PRO-ACT) Database. The data available in the PRO-ACT Database have been volunteered by PRO-ACT Consortium members.

## Author contributions

Y.Y. and H.N. conceived the research project and wrote a manuscript. Y.Y. developed the model and the analytical framework and implemented analysis.

