## Supplemental Information for "Decomposing Heterogeneity in Disease Progression Speeds and Pathways"

Affiliations:

*Corresponding author: Yuichiro Yada

*Co-Corresponding author: Honda Naoki

**Supplementary Table**

Supplementary Table 1: Basic characteristics of the cohort used in the present study

Only patients in each cohort who had records of visiting the hospital at least four times for ALSFRS-R assessments and whose onset location was recorded as limb-onset were included. Patients without recorded covariate values were excluded from the analyses of association with estimated progression speed and from the prognostic prediction. The values for ALSFRS-R Visit Times and Age At Symptom Onset represent the mean and standard deviation.

| Cohort | Number of patients extracted | ALSFRS-R Visit Times | Sex | Age At Symptom Onset | Riluzole Use History |
| --- | --- | --- | --- | --- | --- |
| AnswerALS | 264 | 5.78±1.72 | Female: 86  Male: 177  Not specified: 1 | 55.3±10.9 | TRUE: 177  FALSE: 87 |
| PRO-ACT | 2,565 | 8.88±3.58 | Female: 860  Male: 1705 | 53.7±11.2 | TRUE:1944  FALSE: 621 |

**Supplementary Figures**

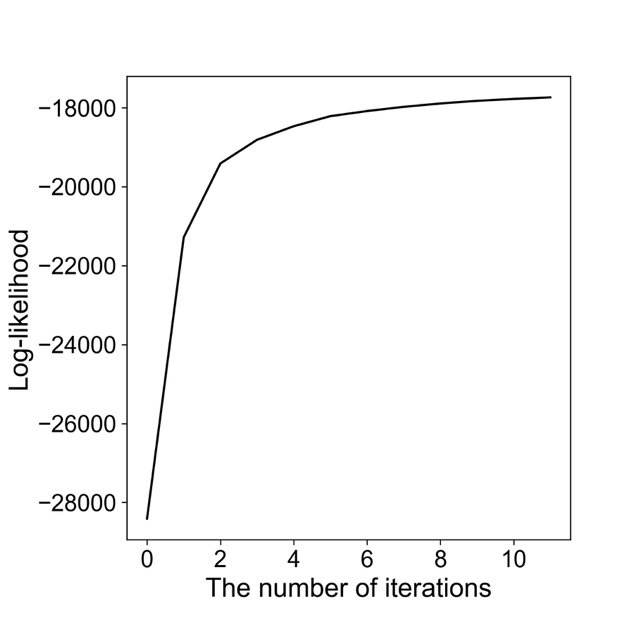

Supplementary Fig. 1: Convergence of model parameter update with EM algorithm

The model parameters of IPS-CT-HMM were updated using the EM algorithm to maximize the likelihood. The maximum number of updates was set to 20, and the updates were terminated when the increase in likelihood due to parameter updates fell below 0.1%.

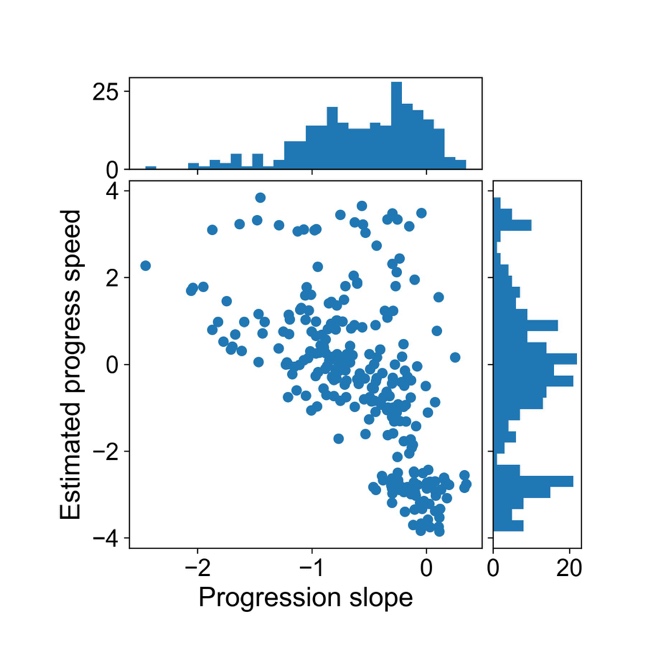

Supplementary Fig. 2: Relationship between the slope of the regression line for the ALSFRS-R total score and the estimated disease progress speed in the AnswerALS cohort
Scatter plot showing the estimated progression speed for patients with a given slope of the regression line for their ALSFRS-R total score. Among patients with a gentle slope, some are estimated to have a faster disease progression.

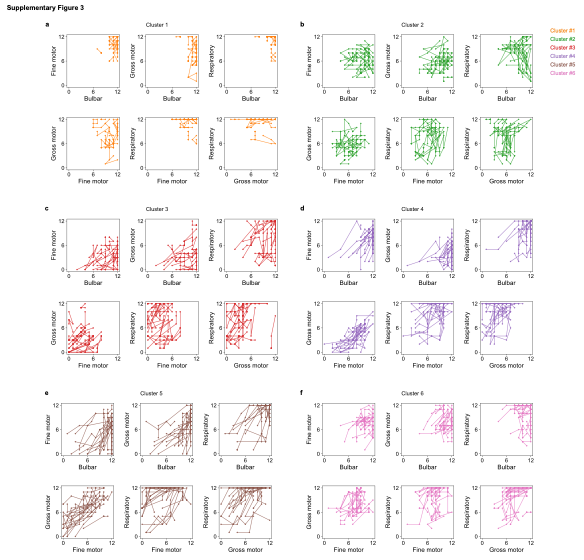

Supplementary Fig. 3: Detailed results of clustering the AnswerALS cohort data.

**a-f,** Figure showing the relationship between functional decline in domains among patients in each cluster identified in the AnswerALS cohort. Subtotals were calculated for each of the ALSFRS-R (Bulbar, Fine motor, Gross motor, Respiratory), and the subtotals for different domains at the same time point are shown.

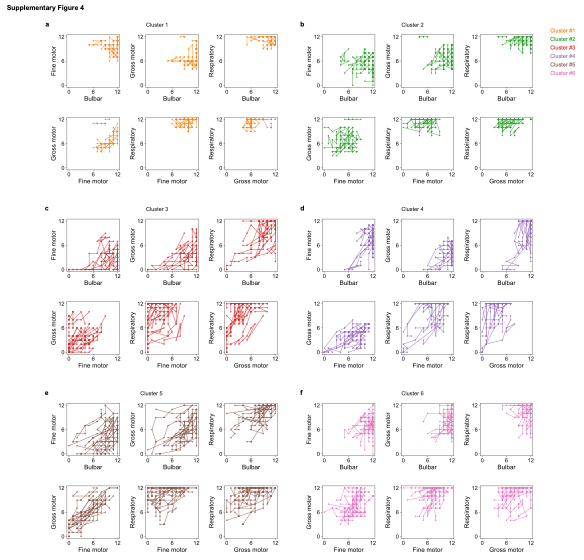

Supplementary Fig. 4: Detailed results of clustering the PRO-ACT cohort data.

**a-f,** Figure showing the classification of PRO-ACT cohort patients and the relationship between the degree of decline in functional domains among patients in each cluster. For classification, each cluster identified in the AnswerALS cohort was referenced. Subtotals were calculated for each domain of the ALSFRS-R (bulbar, fine motor, gross motor, respiratory), and the subtotals for different domains at the same time point are shown.

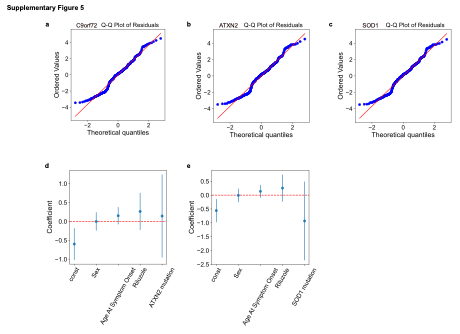

Supplementary Fig. 5: ALS-related mutations, SOD1 and ATXN2, did not showed significant effects on the estimated disease progress speed.

**a-c**, Q–Q plots of standardized residuals from multivariate regression with estimated progression speed as the target variable. The models include **a** C9orf72, **b** ATXN2, or **c** SOD1 genetic predictors. Blue circles show ordered residuals; the red line indicates theoretical normal quantiles. Approximate linearity supports the Gaussian error assumption. **d-e,** Point estimates (dots) and 95% confidence intervals (bars) for coefficients of the multivariate regression with estimated progression speed as the target variable. The models include **d** the ATXN2 mutation term or **e** the SOD1 mutation term as a primary explanatory variable, adjusted for sex, age at symptom onset, and riluzole use. “const” denotes the model intercept and is not interpreted.

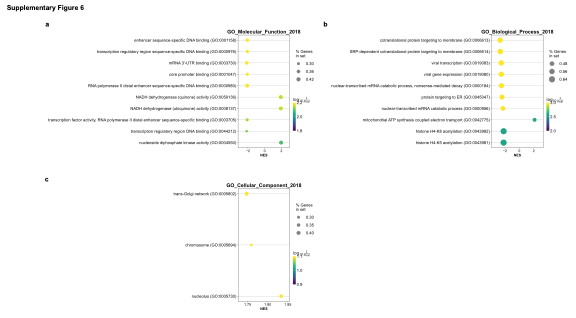

Supplementary Fig. 6: Comprehensive results of gene enrichment analysis on the association between gene/protein expression in patient-derived iPSCs-based motor neurons and estimated disease progress speed.

**a-b,** Extended examples of gene groups exhibiting significant fluctuations in relation to estimated progression speed, obtained from GSEA analysis in the transcriptome. **c**, Those obtained from the proteome.

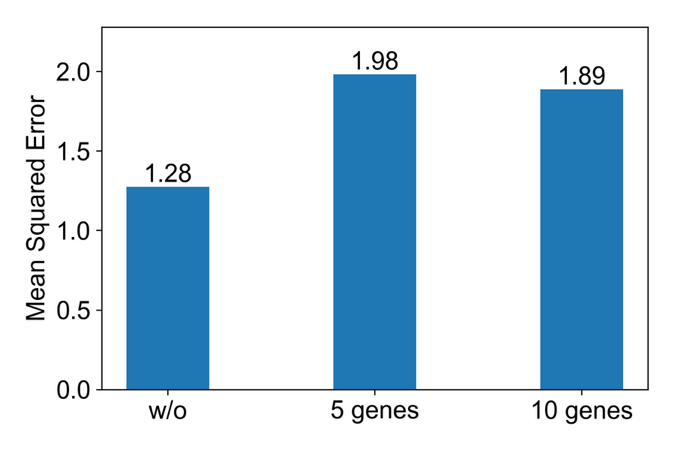

Supplementary Fig. 7: Prediction using transcriptome data obtained from the patient motor neurons in addition to clinical data.

Based on the results of the association analysis between gene expression in patient iPS cell-derived motor neurons and estimated disease progression speeds, genes were sorted in ascending order of p-values. The expression levels of the top 5 or 10 genes, ranked by p-value, were used alongside available clinical information at the start of follow-up to predict estimated progression speeds. The mean squared error was compared between predictions using only clinical information (w/o) and those using both clinical information and gene expression levels (5 genes / 10 genes).
